# Comparative effectiveness of non-pharmacological interventions for primary dysmenorrhea: A protocol for a systematic review and network meta-analysis

**DOI:** 10.1101/2024.03.30.24305112

**Authors:** Qiong-Nan Bao, Jin Yao, Ya-Qin Li, Xin-Yue Zhang, Zheng-Hong Chen, Man-Ze Xia, Wan-Qi Zhong, Ke-Xin Wu, Zi-Han Yin, Fan-Rong Liang

## Abstract

**Introduction:** Primary dysmenorrhea (PD) is the most common gynecological condition among young women. Although several non-pharmacological interventions have proven effective in relieving pain in patients with PD, the optimal treatment remains unknown. This systematic review and network meta-analysis (NMA) will aim to compare and rank the analgesic effects of different non-drug interventions for PD.

**Methods and analysis:** Randomized controlled trials of non-pharmacological interventions for PD will be identified via a search of the PubMed, Cochrane Library, Web of Science, and Embase databases until May 2024. The primary outcome will be change in pain intensity among patients with PD, while the secondary outcomes include health-related quality of life and symptoms of depression and anxiety. Two independent reviewers will perform document screening, study selection, and data extraction. The methodological quality of the included studies will be assessed using the Cochrane Risk of Bias tool (V. 2). The RevMan, Stata, and Aggregate Data Drug Information System software will be used to perform a pairwise meta-analysis and Bayesian NMA in a random-effects model. The certainty of the evidence will be rated using the Grading of Recommendations, Assessment, Development, and Evaluation System.

**Ethics and dissemination:** Ethical approval will not be required for this study because all data will be obtained from published research. The findings will be published in a peer-reviewed journal.

**STRENGTHS AND LIMITATIONS OF THIS STUDY:**

- This will be the first study to comprehensively compare the efficacy of non-pharmacological interventions for primary dysmenorrhea using network meta-analysis.
- The study will assess both short- and long-term efficacies of various non-pharmacological interventions in mitigating pain intensity among patients with primary dysmenorrhea.
- This systematic review will be conducted in compliance with the Cochrane Handbook guidelines, which will ensure methodological rigor.
- Only trials from English databases will be included, which may lead to omission of eligible studies in other languages.
- The efficacy of different regimens of the same non-pharmacological intervention will not be investigated in this study.

## BACKGROUND

Primary dysmenorrhea (PD) is one of the most frequent complaints in women of childbearing age and is characterized by cyclic pelvic pain before or during menstruation without organic diseases.^1^ Severe cases are often accompanied by symptoms such as nausea, back pain, and fatigue.^2^ Epidemiological surveys report the prevalence of PD to be 45% to 95%, with 2% to 29% of menstruating women experiencing severe symptoms.^3-4^ Moreover, PD frequently progresses to chronic pain and significantly impacts the daily lives and psychological states of patients to varying extents,^5-6^ including problems with work or social activities, stress, anxiety, and depression. PD has been reported as the leading cause of school absenteeism in adolescent girls. A recent review estimated that, in the US alone, PD contributes to the loss of 600 million working hours and $2 billion annually due to decreased productivity and increased healthcare costs.^7^ Thus, exploring effective and proper management strategies for PD will be beneficial to both individuals and society.

The exact pathogenesis of PD is not yet fully clear; however, it is widely accepted that this condition is attributed to excessive synthesis and release of prostaglandins that leads to uterine hypercontraction, ischemia, hypoxia along with inflammation and decreased pain threshold. Conventional drug treatments for PD include non-steroidal anti-inflammatory drugs (NSAIDs) and oral contraceptives.^8^ While NSAIDs are highly prescribed to reduce acute pain, long-term use may result in gastrointestinal tract bleeding, cardiovascular risks, and liver/renal complications.^9-10^. Additionally, some women with PD do not respond adequately to NSAIDs.^11^ Oral contraceptives work by suppressing ovulation, but are unsuitable for women who wish to become pregnant or have contraindications. Moreover, they may increase the risk of breast cancer and venous thrombosis.^2 12^ The side effects of these drugs and the nature of the periodic recurrence of menstrual cramps indicate that women are more willing to seek non-pharmacological methods to manage PD.^13^

Currently, a range of non-drug interventions are available for the treatment of PD, including acupuncture (e.g., manual acupuncture, electroacupuncture, auriculotherapy)^14^; transcutaneous electrical nerve stimulation^15^; acupressure^16^; aromatherapy^17^; physical activities (e.g., aerobic exercise, resistance training, balance training, stretching)^18^; physiotherapy (e.g., thermotherapy, electrotherapy, kinesio tape)^19^; psychological intervention (e.g., biofeedback, relaxation, hypnotherapy, imagery)^20^; nutritional and dietary supplements (e.g., vitamin, fennel, zinc, ginger)^21^; and manual therapy (e.g., massage, tuina, manipulation techniques)^22^. Clinical practice guidelines have recommended some of these non-pharmacological interventions for pain relief in PD,^23^ and systematic reviews based on randomized controlled trials (RCTs) have also confirmed that several non-drug therapies can effectively relieve menstrual pain and improve the quality of life (QoL) and emotional well-being in women with PD.^17 19-20 24-27^ Furthermore, these non-pharmacological interventions can not only have a rapid onset but also last for more than one menstrual cycle. For example, acupressure and acupuncture have been reported to significantly reduce pain intensity in patients with PD at both the immediate post-intervention and several months follow-up, compared to baseline.^28 29^ However, the efficacy of distinct non-pharmacological interventions varies, and the optimal regimen for PD management remains unclear, making it difficult for patients and clinicians to select the most appropriate intervention.

Network meta-analysis (NMA) is an analytical method that simultaneously estimates the relative efficacy of multiple treatments by gathering direct and indirect evidence.^30^ To the best of our knowledge, no NMA has proven which non-pharmacological intervention is best for PD. Therefore, we propose this systematic review and NMA protocol to compare and rank the short-term and long-term analgesic effects of non-pharmacological interventions in patients with PD. The findings of this study will provide a reference to inform recommendations for decision makers in clinical and policy settings.

## METHODS

### Study registration

This protocol has been registered at PROSPERO (CRD42023459026) and designed and written according to the Preferred Reporting Items for Systematic Reviews and Meta-Analyses Protocol (PRISMA-P) statement.^32^ The NMA will be performed in compliance with the guidelines of the Cochrane Handbook,^32^ and findings will be reported following the guidelines of the PRISMA-NMA.^33^

### Eligiblity criteria

#### Types of studies

Only RCTs published in English will be included in the meta-analysis. Non-RCTs, articles with unavailable full texts, and animal studies will be excluded.

#### Types of participants

Studies that enrolled women who had menstrual pain without underlying causes with regular menstrual periods and cycles will be included, regardless of age, severity, or disease duration. Studies including women with secondary dysmenorrhea will be excluded.

#### Types of interventions

Non-pharmacological interventions, encompassing a range of modalities such as acupuncture therapy,^34^ acupressure,^35^ aromatherapy,^17^ exercise,^23^ TENS,^36^ physiotherapy,^37^ psychological intervention,^20^ manual therapy,^38^ moxibustion,^39^ nutritional therapy,^25^ non-invasive brain stimulation, used as treatment will be included.^40^ No restrictions will be placed on the intervention timing (pre-menstrual, menstrual period, post-menstrual), courses, or delivery form (researchers, self, or device). Interventions involving pharmacology, including analgesics, contraceptives, and other drugs,will be excluded. Studies where participants were permitted to take necessary medication in the event of intolerable pain will be included.

#### Types of control groups

Control groups that received another of the non-pharmacological interventions mentioned above, no treatment, or placebo or placed on a waiting-list, will be eligible for study enrolment. Studies that used drugs or distinctive types of the same treatment as the controls will be excluded.

#### Types of outcome measures

##### Primary outcomes

The primary outcomes will be the short- and long-term efficacy of non-pharmacological treatment. Short-term efficacy has been defined as changes in pain intensity from baseline to the end of treatment. Long-term efficacy has been defined as changes in pain intensity from baseline to the end of follow-up (1-12 months).^41^ Included studies will evaluate pain intensity using standardized and validated scales, including the visual analog scale and numeric rating scale.^42^

##### Secondary outcomes

Changes in health-related QoL and psychological problems (depression and anxiety) from baseline to post-intervention will be considered secondary outcomes. Included studies will have used the Short Form 36, Self Rating Anxiety Scale, and Self Rating Depression Scale to evaluate QoL, anxiety, and depression, respectively.

### Data sources and search strategy

PubMed, Cochrane Library, Web of Science, and Embase databases will be searched for articles published between January 2000 and May 2024. Clinical registry platform (Clinical trials.gov and ChiCTR) will also be retrieved for unpublished trials.

Additionally, the reference lists of the included papers and relevant reviews will be manually searched to identify qualified studies. The search will be limited to English-language publications. The search terms will include “randomized controlled trial”, “dysmenorrhea”, “acupuncture”, “acupressure”, “aromatherapy”, “exercise”, “nerve stimulation”, “brain stimulation”, “massage”, “moxibustion”, “physiotherapy”, “thermotherapy”, “electrotherapy”, “manual therapy”, “hypnotherapy”, “biofeedback”, “relaxation”, “nutrition”, “ginger”, etc. The detailed search strategy for each database is presented in Supplemental Appendix 1.

### Selection of studies

All retrieved records will be imported into Endnote V.X9, and duplicates will be automatically removed. Two authors will independently browse the titles and abstracts for initial eligibility, and the final selection of eligible studies will be based on a full-text review. Any differences between reviewers will be resolved through discussion with an experienced investigator. The PRISMA flowchart of the selection procedure is shown in Figure 1.

**Figure 1.**
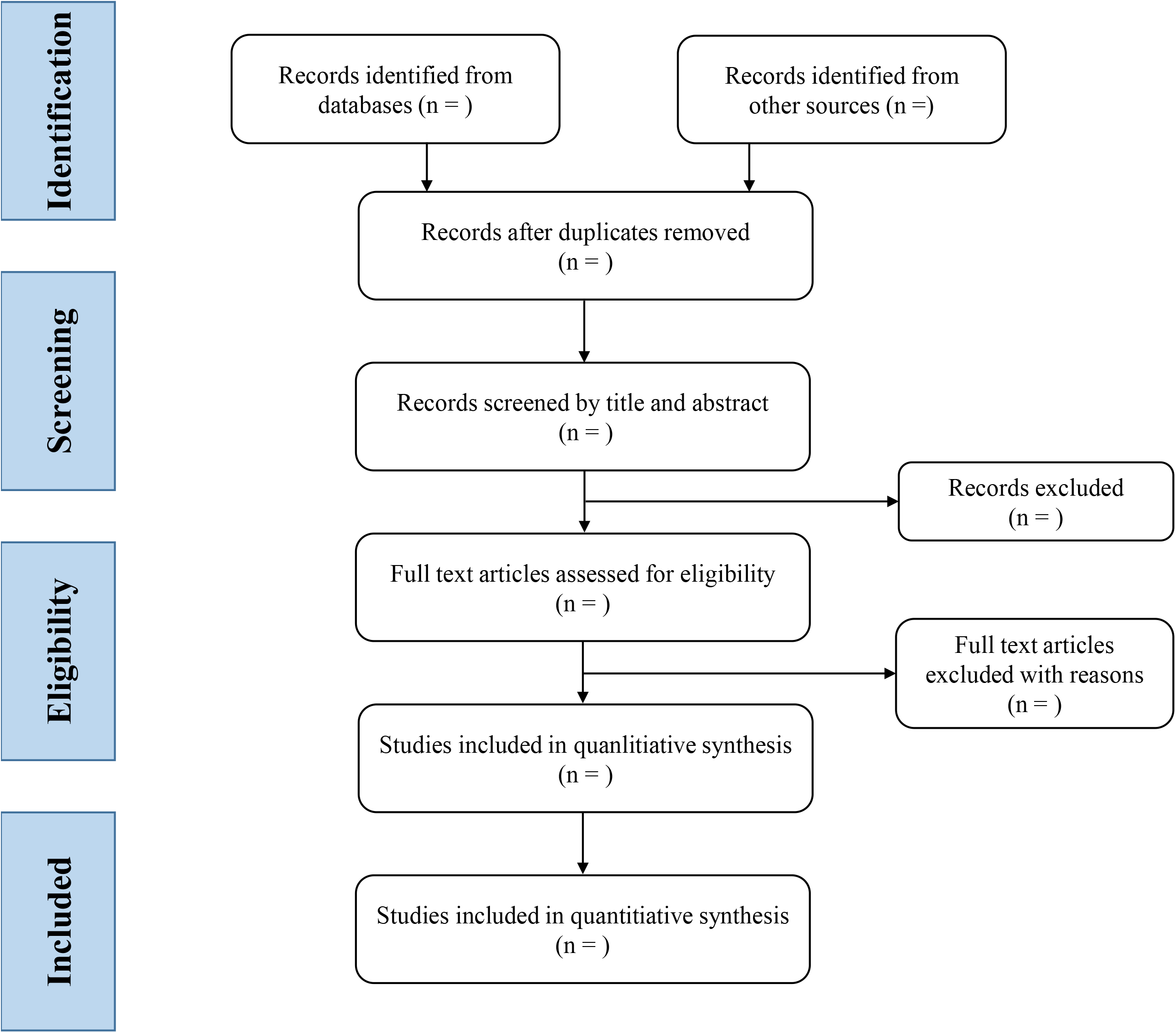
PRISMA flow diagram of the study selection process.

### Data extraction and management

Two reviewers will independently extract essential information from the included studies using a predesigned Excel spreadsheet. The extracted data will include details, such as the first author, publication year, country, age, sample size, intervention and control methods, pain intensity before treatment, treatment session and duration, follow-up period, time points, outcome measures data, and adverse events. If pain intensity in multiple regions (e.g., lower abdominal pain, back pain, and thigh pain) were measured in a study, the outcome of lower abdominal pain will be extracted. In studies where multiple follow-up time points have been reported, the outcome of the longest time point will be extracted. If multiple pain intensities (e.g., peak and average pain intensity) were reported, we will extract data on the average pain intensity. If pain intensity was assessed on multiple days of one menstrual cycle, we will extract the results of the first day of the menstrual cycle when dysmenorrhea frequently initiates.^43^ For missing data or unclear information that could affect the analysis, the corresponding author will be contacted to acquire as much detailed information as possible.

### Risk of bias assessment

The methodological quality of all included studies will be assessed by two independent reviewers using the Cochrane Risk-of-Bias (RoB) 2.0 tool.^44^ The overall RoB will be evaluated as low, high, or with some concerns across various domains, including randomization process, deviations from intended interventions, missing outcome data, outcome measurement, and selective reporting of results. Any disputes during the assessment will be resolved by a third reviewer.

### Statistical analysis

#### Pairwise meta-analysis

We will perform a pairwise meta-analysis using RevMan software V.5.4 (Review Manager, The Cochrane Collaboration, 2020) to compare the interventions with direct evidence. All data will be continuous variables, and the mean difference (MD) with 95% credible intervals (CIs) will be synthesized as a relative treatment effect. If mean and SD are not available, we will calculate them from the reported data statistics (e.g., median and range, SEs, and 95% CIs) according to the corresponding formula. The heterogeneity between direct comparisons will be estimated by using *I*^2^ statistics and *P* value. *I*^2^ values of > 30%, 50%, and 75% represent moderate, substantial, and considerable heterogeneity respectively.^45^ The random-effects model will be adopted in case substantial heterogeneity is observed (*I*^2^ > 50%), and possible sources of heterogeneity will be explored using prespecified subgroup analyses.

#### Network meta-analysis

The NMA will be implemented in a Bayesian random-effects framework using the Aggregate Data Drug Information System (ADDIS) software V.1.16.6 (Drugis, Groningen, Netherlands). The Markov chain Monte Carlo method will be used for all analyses, the parameters include four chains with 50000 simulation iterations, the initial burn-in of 10000 will be discarded. The node-splitting method will be used to examine the inconsistency between direct and indirect comparisons. The inconsistency model will be chosen when *P* < 0.05, which represents a statistically significant difference; otherwise, the consistency model will be selected. The potential scale reduction factor (PSRF) value will be analysed to evaluate the convergence of the pooled results. A PSRF value close to 1 indicates successful convergence.

Network plots will be created using the Stata software V.15.1 (Stata Corp, College Station, Texas, USA) to present the geometry of all comparisons in the included studies. The nodes in these plots signify different non-pharmacological interventions, and the lines between every pair of nodes imply a direct comparison. Larger nodes represent a larger sample size, and thicker lines reflect a greater number of RCTs.

Finally, ranking probability plots will be generated for each intervention.

### Subgroup analysis and sensitivity analysis

If sufficient data are available, we plan to perform the subgroup analyses based on different intervention timing (e.g., premenstrual, menstrual period), and treatment courses. Sensitivity analysis will be performed to verify the stability of our results by excluding trials with a high risk of bias, a dropout rate of more than 20 %, and small sample sizes.

### Publication bias assessment

If 10 or more trials are included, a comparison-adjusted funnel plot^46^ will be generated to evaluate the reporting bias. If the funnel plot is asymmetrical, a publication bias may exist.

### Evidence quality assessment

The Grading of Recommendations, Assessment, Development, and Evaluation (GRADE) Profiler software will be applied to evaluate the strength of the evidence for all outcomes. The quality of evidence will be judged as “high,” “moderate,” “low,” or “very low” quality in terms of the down-graded factors (risk of bias, inconsistency, indirectness, imprecision, and publication bias) and up-graded factors (large effect, dose-response, all plausible confounding).^47^

### Patient and public involvement

None.

## DISCUSSION

PD is prevalent in reproductive women.^48^ Due to recurrent monthly pain, patients with PD commonly experience decreased QoL and poor physical and mental health, which may, in turn, worsen the severity of pain.^13 49^ Currently, various non-pharmacological interventions are used to manage menstrual pain and its associated symptoms in women with PD. However, it is unknown which method works best for pain relief.

Therefore, it is important to compare the efficacy of non-pharmacological interventions in patients with PD using NMA.^50^ Our findings will provide a ranking of non-drug interventions for PD to assist patients and therapists in decision-making.

This study has several strengths: First, it will be the first and most extensive NMA to verify the analgesic effectiveness of drug-free interventions for PD and determine the best intervention. Second, we will include as many non-drug therapies as possible, since the planned literature search strategy is wide and representative. Third, we will investigate the comparative effects of various non-drug interventions on the QoL and emotional status of women with PD.

However, there will be certain unavoidable limitations as well. First, heterogeneity between studies may be high due to the variability of various methods; however, we intend to explore any sources of heterogeneity through sub-group analysis. Lastly, including only trials published in English may result in bias.

## Data Availability

All data produced in the present study are available upon reasonable request to the authors

## Author’s contribution

QNB conceptualized this study, QNB, and JY wrote the first draft of the current protocol, with ZHY and FRL critically revised the final manuscript. YQL and XYZ developed the protocol methodology. Literature searching, screening, data extraction, analyses, and methodological assessment will be implemented by WQZ, KXW, ZHC and MZX. Z-HY will arbitrate any conflicts between reviewers and perform quality checks at all stages of review. All authors read, provided feedback, and approved the submitted version.

