## Supplementary File 1 for "Comparative effectiveness of non-pharmacological interventions for primary dysmenorrhea: A protocol for a systematic review and network meta-analysis"

Supplementary Appendix 1: Search strategies for each database

PubMed

#1 "dysmenorrhea"[MeSH Terms]

#2 "dysmenorrhea*"[Title/Abstract] OR "menstrual pain*"[Title/Abstract] OR "menstruation pain*"[Title/Abstract] OR "menstrual cramp*"[Title/Abstract] OR "period pain*"[Title/Abstract] OR "painful menstruation*"[Title/Abstract] OR "pain, menstrual"[Title/Abstract]

#3 #1 OR #2

#4 "Acupuncture Therapy"[MeSH Terms] OR "Electroacupuncture"[MeSH Terms] OR "Auriculotherapy"[MeSH Terms] OR "Acupressure"[MeSH Terms] OR "Exercise Therapy"[MeSH Terms] OR "Transcutaneous Electric Nerve Stimulation"[MeSH Terms] OR "Aromatherapy"[MeSH Terms] OR "Moxibustion"[MeSH Terms] OR "Massage"[MeSH Terms] OR "Yoga"[MeSH Terms] OR "Dance Therapy"[MeSH Terms] OR "Electric Stimulation Therapy"[MeSH Terms] OR "Biofeedback, Psychology"[MeSH Terms] OR "Hypnosis"[MeSH Terms] OR "Ginger"[MeSH Terms] OR "Relaxation Therapy"[MeSH Terms] OR "Behavior Therapy"[MeSH Terms] OR "Musculoskeletal Manipulations"[MeSH Terms]

#5 "nonpharmacological"[Title/Abstract] OR "nondrug"[Title/Abstract] OR "acupuncture"[Title/Abstract] OR "electroacupuncture"[Title/Abstract] OR "acupressure"[Title/Abstract] OR "acupoint*"[Title/Abstract] OR "auricular"[Title/Abstract] OR "auriculotherapy"[Title/Abstract] OR "nerve stimulation"[Title/Abstract] OR "neurostimulation"[Title/Abstract] OR "electrical stimulation"[Title/Abstract] OR "current stimulation"[Title/Abstract] OR "massage"[Title/Abstract] OR "tuina"[Title/Abstract] OR "dry needl*"[Title/Abstract] OR "moxibustion"[Title/Abstract] OR "exercise*"[Title/Abstract] OR "yoga"[Title/Abstract] OR "dance"[Title/Abstract] OR "pilates"[Title/Abstract] OR "physical activity"[Title/Abstract] OR "physiotherapy"[Title/Abstract] OR "physical therapy"[Title/Abstract] OR "topical heat"[Title/Abstract] OR "local heat"[Title/Abstract] OR "heat therapy"[Title/Abstract] OR "thermotherapy"[Title/Abstract] OR "electrotherapy"[Title/Abstract] OR "biofeedback"[Title/Abstract] OR "hypnotherapy"[Title/Abstract] OR "ginger"[Title/Abstract] OR "relaxation"[Title/Abstract] OR "behavior* therapy"[Title/Abstract] OR "manipulative therapy"[Title/Abstract] OR "manual therapy"[Title/Abstract] OR "manipulation"[Title/Abstract] OR "kinesio tape"[Title/Abstract]

#6 #4 OR #5

#7 "Randomized Controlled Trials as Topic"[MeSH Terms] OR "random allocation"[MeSH Terms] OR "randomized controlled trial"[Publication Type] OR "controlled clinical trial"[Publication Type] OR "clinical trial"[Publication Type] OR "clinical study"[Title/Abstract] OR "trial"[Title/Abstract] OR "placebo"[Title/Abstract] OR "random*"[Title/Abstract]

#8 #3 AND #6 AND #7

EMBASE

#1 ‘dysmenorrhea’/exp/mj OR ‘dysmenorrhea’:ti,ab,kw OR ‘menstrual pain’:ti,ab,kw OR ‘menstruation pain’:ti,ab,kw OR ‘menstrual cramp’:ti,ab,kw OR ‘period pain’:ti,ab,kw OR ‘painful menstruation’:ti,ab,kw OR ‘pain, menstrual’:ti,ab,kw

#2 ‘acupuncture therapy’/exp/mj OR ‘electroacupuncture’/exp/mj OR ‘exercise therapy’/exp/mj OR ‘acupressure’/exp/mj OR ‘auriculotherapy’/exp/mj OR ‘transcutaneous electric nerve stimulation’/exp/mj OR ‘aromatherapy’/exp/mj OR ‘moxibustion’/exp/mj OR ‘massage’/exp/mj OR ‘yoga’/exp/mj OR ‘Dance Therapy’/exp/mj OR ‘Electric Stimulation Therapy’/exp/mj OR ‘Biofeedback, Psychology’/exp/mj OR ‘Hypnosis’/exp/mj OR ‘Ginger’/exp/mj OR ‘Relaxation Therapy’/exp/mj OR ‘Behavior Therapy’/exp/mj OR ‘Musculoskeletal Manipulations’/exp/mj OR ‘nonpharmacological’:ti,ab,kw OR ‘nondrug’:ti,ab,kw OR ‘acupuncture’:ti,ab,kw OR ‘electroacupuncture’:ti,ab,kw OR ‘acupoint’:ti,ab,kw OR ‘auriculotherapy’:ti,ab,kw OR ‘auricular’:ti,ab,kw OR ‘acupressure’:ti,ab,kw OR ‘dry needl*’:ti,ab,kw OR ‘nerve stimulation’:ti,ab,kw OR ‘electrical stimulation’:ti,ab,kw OR ‘massage’:ti,ab,kw OR ‘moxibustion’:ti,ab,kw OR ‘exercise’:ti,ab,kw OR ‘neurostimulation’:ti,ab,kw OR ‘current stimulation’:ti,ab,kw OR ‘tuina’:ti,ab,kw OR ‘yoga’:ti,ab,kw OR ‘dance’:ti,ab,kw OR ‘pilates’:ti,ab,kw OR ‘physical activity’:ti,ab,kw OR ‘physiotherapy’:ti,ab,kw OR ‘physical therapy’:ti,ab,kw OR ‘topical heat’:ti,ab,kw OR ‘local heat’:ti,ab,kw OR ‘heat therapy’:ti,ab,kw OR ‘thermotherapy’:ti,ab,kw OR ‘electrotherapy’:ti,ab,kw OR ‘biofeedback’:ti,ab,kw OR ‘hypnotherapy’:ti,ab,kw OR ‘ginger’:ti,ab,kw OR ‘relaxation’:ti,ab,kw OR ‘behavior* therapy’:ti,ab,kw OR ‘manipulative therapy’:ti,ab,kw OR ‘manual therapy’:ti,ab,kw OR ‘manipulation’:ti,ab,kw OR ‘kinesio tape’:ti,ab,kw

#3 ‘Randomized Controlled Trials as Topic’/exp/mj OR ‘random allocation’/exp/mj OR ‘clinical study’:ti,ab,kw OR ‘trial’:ti,ab,kw OR ‘placebo’:ti,ab,kw OR ‘randomly’:ti,ab,kw OR ‘randomized’:ti,ab,kw

#4 #1 AND #2 AND #3

Web of science

TS=(‘dysmenorrhea*’ OR ‘menstrual pain*’ OR ‘menstruation pain*’ OR ‘menstrual cramp*’ OR ‘period pain*’ OR ‘painful menstruation*’ OR ‘pain, menstrual’) AND TS=(‘nonpharmacological’ OR ‘nondrug’ OR ‘acupuncture’ OR ‘electroacupuncture’ OR ‘dry needl*’ OR ‘acupoint*’ OR ‘auriculotherapy’ OR ‘auricular’ OR ‘acupressure’ OR ‘nerve stimulation’ OR ‘electrical stimulation’ OR ‘moxibustion’ OR ‘massage’ OR ‘exercise*’ OR ‘tuina’ OR ‘neurostimulation’ OR ‘current stimulation’ OR ‘yoga’ OR ‘dance’ OR ‘pilates’ OR ‘physical activity’ OR ‘physiotherapy’ OR ‘physical therapy’ OR ‘topical heat’ OR ‘local heat’ OR ‘heat therapy’ OR ‘thermotherapy’ OR ‘electrotherapy’ OR ‘biofeedback’ OR ‘hypnotherapy’ OR ‘ginger’ OR ‘relaxation’ OR ‘behavior* therapy’ OR ‘manipulative therapy’ OR ‘manual therapy’ OR ‘manipulation’ OR ‘kinesio tape’) AND TS=(‘Randomized Controlled Trials as Topic’ OR ‘random allocation’ OR ‘clinical study’ OR ‘trial’ OR ‘placebo’ OR ‘random*’)

Cochrane Library

#1 Mesh descriptor: [dysmenorrhea] explode all trees

#2 dysmenorrhea*:ti,ab,kw OR menstrual pain:ti,ab,kw OR menstruation pain:ti,ab,kw OR menstrual cramp:ti,ab,kw OR period pain:ti,ab,kw OR painful menstruation:ti,ab,kw OR pain, menstrual:ti,ab,kw

#3 #1 OR #2

#4 Mesh descriptor: [acupuncture therapy] explode all trees

#5 Mesh descriptor: [electroacupuncture] explode all trees

#6 Mesh descriptor: [acupressure] explode all trees

#7 Mesh descriptor: [auriculotherapy] explode all trees

#8 Mesh descriptor: [transcutaneous electric nerve stimulation] explode all trees

#9 Mesh descriptor: [moxibustion] explode all trees

#10 Mesh descriptor: [exercise therapy] explode all trees

#11 Mesh descriptor: [aromatherapy] explode all trees

#12 Mesh descriptor: [massage] explode all trees

#13 Mesh descriptor: [yoga] explode all trees

#14 Mesh descriptor: [Dance Therapy] explode all trees

#15 Mesh descriptor: [Electric Stimulation Therapy] explode all trees

#16 Mesh descriptor: [Biofeedback, Psychology] explode all trees

#17 Mesh descriptor: [Hypnosis] explode all trees

#18 Mesh descriptor: [Ginger] explode all trees

#19 Mesh descriptor: [Relaxation Therapy] explode all trees

#20Mesh descriptor: [Behavior Therapy] explode all trees

#21 Mesh descriptor: [Musculoskeletal Manipulations] explode all trees

#22 nonpharmacological:ti,ab,kw OR nondrug:ti,ab,kw OR acupuncture:ti,ab,kw OR electroacupuncture:ti,ab,kw OR acupoint:ti,ab,kw OR "dry needl*":ti,ab,kw auriculotherapy:ti,ab,kw OR auricular:ti,ab,kw OR acupressure:ti,ab,kw OR "electrical stimulation":ti,ab,kw OR "nerve stimulation":ti,ab,kw OR massage:ti,ab,kw OR moxibustion:ti,ab,kw OR exercise*:ti,ab,kw OR neurostimulation:ti,ab,kw OR current stimulation:ti,ab,kw OR tuina:ti,ab,kw OR yoga:ti,ab,kw OR dance:ti,ab,kw OR pilates:ti,ab,kw OR physical activity:ti,ab,kw OR physiotherapy:ti,ab,kw OR physical therapy:ti,ab,kw OR topical heat:ti,ab,kw OR local heat:ti,ab,kw OR heat therapy:ti,ab,kw OR thermotherapy:ti,ab,kw OR electrotherapy:ti,ab,kw OR biofeedback:ti,ab,kw OR hypnotherapy:ti,ab,kw OR ginger:ti,ab,kw OR relaxation:ti,ab,kw OR "behavior* therapy":ti,ab,kw OR "manipulative therapy":ti,ab,kw OR "manual therapy":ti,ab,kw OR "manipulation":ti,ab,kw OR "kinesio tape":ti,ab,kw

#23 #4 OR #5 OR #6 OR #7 OR #8 OR #9 OR #10 OR #11 OR #12 OR #13 OR #14 OR #15 OR #16 OR #17 OR #18 OR #19 OR #20 OR #21 OR #22

#24 Mesh descriptor: [Randomized Controlled Trials as Topic] explode all trees

#25 Mesh descriptor: [random allocation] explode all trees

#26 "clinical study":ti,ab,kw OR trial:ti,ab,kw OR placebo:ti,ab,kw OR ranom*:ti,ab,kw

#27 #24 OR #25 OR #26

#28 #3 AND #23 AND #27
